# High-throughput immunoassays for SARS-CoV-2 – considerable differences in performance when comparing three methods

**DOI:** 10.1101/2020.05.22.20106294

**Authors:** Oskar Ekelund, Kim Ekblom, Sofia Somajo, Johanna Pattison-Granberg, Karl Olsson, Annika Petersson

## Abstract

**Background:** The recently launched high-throughput assays for the detection of antibodies against SARS-CoV-2 may change the managing strategies for the COVID-19 pandemic. This study aimed at investigating the performance of three high-throughput assays and one rapid lateral flow test relative to the recommended criteria defined by regulatory authorities.

**Methods:** A total of 133 samples, including 100 pre-pandemic samples, 20 samples from SARS-CoV-2 RT-PCR positive individuals, and 13 potentially cross-reactive samples were analysed with SARS-CoV-2 IgG (Abbott), Elecsys Anti-SARS-CoV-2 (Roche), LIAISON SARS-CoV-2 S1/S2 IgG (DiaSorin) and 2019-nCOV IgG/IgM Rapid Test (Dynamiker Biotechnology Co).

**Results:** All assays performed with a high level of specificity; however, only Abbott reached 100% (95% CI 96.3-100). The pre-pandemic samples analysed with Roche, DiaSorin and Dynamiker Biotechnology resulted in two to three false-positive results per method (specificity 96.9-98.0%). Sensitivity differed more between the assays, Roche exhibiting the highest sensitivity (100%, CI 83.9-100). The corresponding figures for Abbott, DiaSorin and Dynamiker Biotechnology were 85.0%, 77.8% and 75.0%, respectively.

**Conclusions:** The results of the evaluated SARS-CoV-2 assays vary considerably as well as their ability to fulfil the performance criteria proposed by regulatory authorities. Introduction into clinical use in low-prevalent settings, should therefore, be made with caution.

## Introduction

Coronavirus disease 2019 (COVID-19) was first reported in Wuhan, Hubei province, China, in December 2019 [1]. The virus causing this disease has since been designated SARS-CoV-2 [2]. The clinical manifestations of COVID-19 range from mild or no respiratory symptoms to severe viral pneumonia with a need of intensive care and ventilator support. Due to the rapid dissemination of the infection, the outbreak was declared a pandemic by the World Health Organization (WHO) on 11^th^ March 2020 [3].

RT-PCR detection of SARS-CoV-2 in samples primarily obtained from nasopharyngeal and/or pharyngeal swabs is considered gold standard for laboratory confirmation of COVID-19 in patients. However, the presence of viral nucleic acids is transient, and results are dependent on the time of sampling.

Detection of antibodies against the SARS-CoV-2 virus could possibly be used to distinguish patients recovered from COVID-19, and several rapid tests based on immunochromatographic techniques have been developed. These point-of-care tests usually deliver results within 15-30 minutes, however the nature of the tests makes large-scale testing inefficient.

To achieve high-throughput detection of anti-SARS-CoV-2 antibodies in plasma or serum, several manufacturers have developed immunoassays based on recognition of recombinant antigens. Some tests claim to detect IgM or IgG, while others also identify IgA or a mixture of different classes of antibodies. Differences in test design, including choice of antigen, are likely to affect the sensitivity and specificity of the tests [4]. In this study, the performance of three commercially available high-throughput automated SARS-CoV-2 antibody assays, and one rapid immunochromatographic test, was investigated and compared to recommended criteria set by regulatory authorities.

## Materials and Methods

### Assays

Four commercially available CE marked immunoassays, and their corresponding platforms were used: 1) Abbott SARS-CoV-2 IgG on the ARCHITECT i2000 (Abbott, Illinois, USA); 2) Elecsys Anti-SARS-CoV-2 on the Cobas 8000 e801 (Roche Diagnostic Scandinavia AB, Solna, Sweden); 3) LIAISON SARS-CoV-2 S1/S2 IgG on the LIAISON XL (DiaSorin, Saluggia, Italy); and 4) the lateral flow test 2019-nCOV IgG/IgM Rapid Test (Dynamiker Biotechnology Co., Tianjin, China).

The Abbott SARS-CoV-2 IgG reagent is based on the recognition of IgG antibodies binding to recombinant SARS-CoV-2 nucleoprotein in a chemiluminescent microparticle immunoassay (CMIA). Results are obtained after 29 minutes, and are reported as an index with a cut-off to distinguish between negative and positive results.

The Elecsys Anti-SARS-CoV-2 assay is based on recombinantly produced SARS-CoV-2 nucleocapsid (N) protein and detects antibodies (including IgG). The duration of the analysis is approximately 18 minutes, with results presented as a cut-off index as well as reactive or non-reactive.

The LIAISON SARS-CoV-2 S1/S2 detects IgG antibodies recognising the spike glycoprotein of the coronavirus. As for the previous assays, the antigen is composed of recombinant protein expressed in human cell lines. The analysis takes 35 minutes, and the results are expressed as IgG antibody concentrations in arbitrary units (AU/mL) graded negative, equivocal, or positive.

The manually performed 2019-nCOV IgG/IgM Rapid Test recognises and differentiates IgG from IgM antibodies; however, information on antigen source is scarce. The test result could be obtained after approximately 10 minutes as visual bands across the assay paper strip, any positive result, either for IgG, IgM, or both, was considered positive. Despite the short reaction time, as compared with the automatic tests, high-throughput analysis of a large number of samples using the manual test procedure is implausible.

All automated systems are part of the routine operations in our laboratories and were as such subjected to accepted quality assurance procedures. All tests (including calibration and controls) were performed according to manufacturers’ instructions, using serum samples.

### Sample collections

All samples in the study originated from an existing sample collection at the Microbiology department obtained after consent to deposit, store, and use for research and development. Samples were fully anonymised prior to inclusion, and results could hence not be linked to individuals. As a consequence, the study did not require approval from an ethics committee, according to the guidelines of the Swedish Ethical Review Agency. All serum samples were stored at −20°C until analysis.

The specificity of each assay was evaluated using 100 pre-pandemic (2018) samples. To challenge the assays, 13 additional serum samples with possible interferences (antinuclear antibodies (n=2); rheumatoid factor (n=2); anti-cytomegalovirus IgM (n=2); anti-Epstein-Barr virus IgM (n=2) and samples from pregnant donors (n=5)) were analysed.

Assay sensitivities were evaluated using 20 outpatient serum samples from 16 individuals that prior to serum sampling had tested RT-PCR positive for SARS-CoV-2 from nasopharyngeal and/or pharyngeal swabs. RT-PCR had been performed using primers and probes targeting either the envelope (E) and the polymerase (RdRP) genes of SARS-CoV-2 as described by Corman [5], or the nucleocapsid gene (N) of the virus (Abbott RealTime SARS-CoV-2 Assay, Abbott Molecular Inc., Illinois, USA). The interval between onset of COVID-19 symptoms and serum sample collection ranged from 18 to 52 days (median 38 days).

### Calculations

Overall per cent agreement, sensitivity (per cent positive agreement), and specificity (per cent negative agreement) were calculated based on a contingency table according to EP12-A2 [6], using Microsoft Excel 2019 (Microsoft Corp. Redmond, WA, USA). The between-test agreement was evaluated using Cohen’s kappa calculated with IBM SPSS Statistics for Windows, Version 26.0 (IBM Corp., Armonk, NY, USA). For each assay, the positive likelihood ratio (LR+) and positive predictive value (PPV) at different prevalence (P) scenarios were calculated according to equation 1 and 2, respectively.

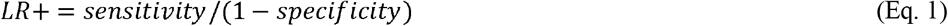

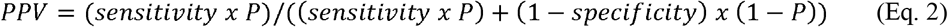

Further data analysis, including descriptive statistics, was performed using GraphPad Prism, version 7.04 for Windows (GraphPad Software, La Jolla California USA, www.graphpad.com).

### Clinical Performance Requirements

The performance of each assay was compared with published guidelines from three regulatory authorities: the Public Health Agency of Sweden (PHAS); Haute Autorité de Santé, France (HAS); and Centres for Disease Control and Prevention, USA (CDC) [7-9]. Their recommended performance criteria are summarized in Table 1.

**Table 1.** Summary of recommended criteria for SARS-CoV-2 serology assays issued by authorities in three different countries.

| Regulatory Authority | Sensitivity (%) | Specificity (%) | Additional comment |
| --- | --- | --- | --- |
| Public Health Agency of Sweden | 90.0 | 99.5 | Recommendations based on a seroprevalence of 5%, rendering a target PPV of > 90% |
| Haute Autorité de Santé, France | 95.0 | 98.0 |  |
| Centers for Disease Control and Prevention, USA | - | 99.5 | For populations with seroprevalence of $\geq 5\%$ |

## Results

Of the 100 pre-pandemic samples, nine tested false positive, two with Roche, five with DiaSorin (including two equivocal results) and two with Dynamiker Biotechnology (figure 1). The negative sample collection resulted in median values of 0.09 COI (0.08-3.16), 0.04 S/CO (0.01-0.6) and 5.7 AU/mL (1.9-32.6) for Roche, Abbott and DiaSorin, respectively. The panel consisting of 13 potentially cross-reactive pre-pandemic samples mainly gave negative results. However, for Dynamiker Biotechnology, one sample with rheumatoid factor IgM was positive for both SARS-CoV-2 IgM and IgG. Also, one sample obtained during pregnancy showed an equivocal result on the DiaSorin assay. In contrast, all these samples were negative on the Abbott and Roche assays.

Three RT-PCR positive samples gave a weak positive result with Roche, close to the cut-off (1.01 to 1.26, cut-off ≥1.0) (Figure 1). These samples were reported as negative by DiaSorin, Dynamiker Biotechnology, and Abbott. However, for these samples, the Abbott values clearly differed from the rest of the Abbott negative samples by being close to the cut-off (0.81-1.06, cut-off ≥1.4).

**Figure 1.**
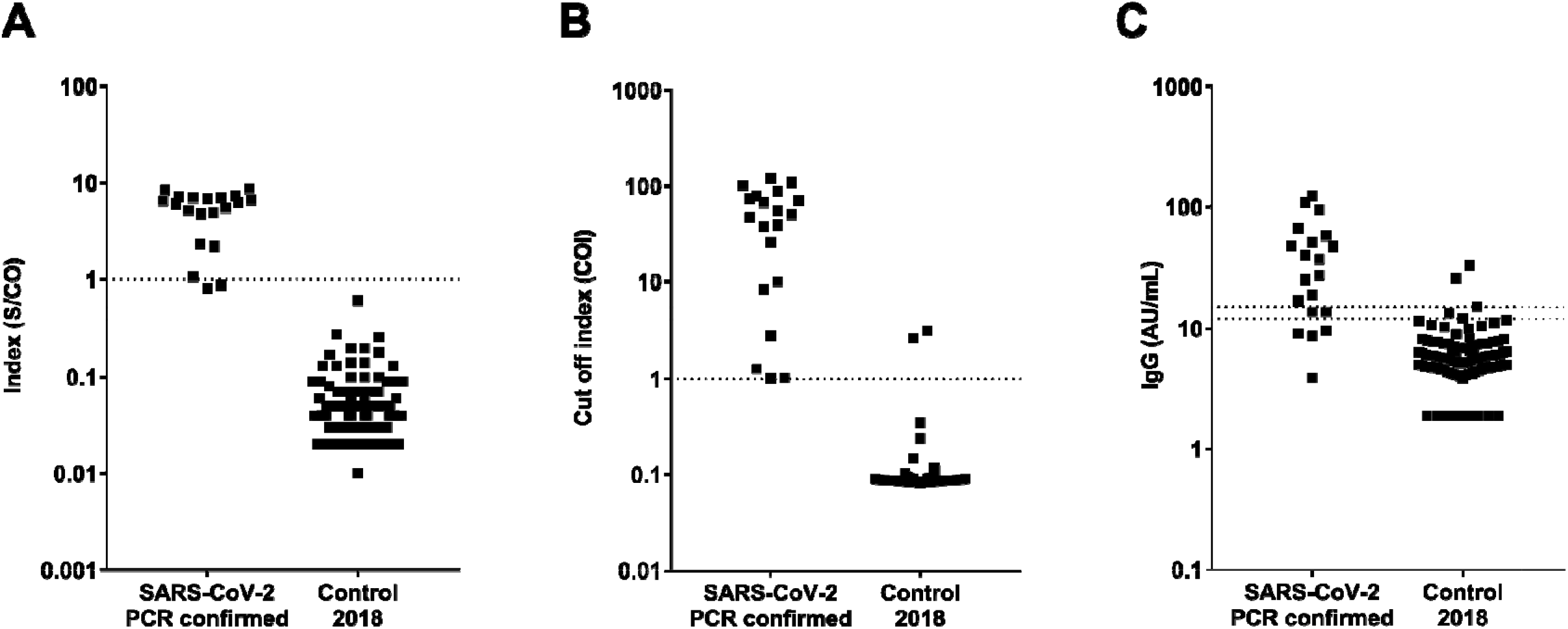
Differences in distribution patterns between Abbott, Roche and DiaSorin immunoassays for detection of SARS-CoV-2 antibodies. Measured values from serology testing of 20 positive (SARS-CoV-2 PCR confirmed) and 100 negative (pre-pandemic 2018) samples. Dotted lines represent cut-off values of A) Abbott: positive result index ≥1.4 S/CO, B) Roche: positive cut-off index (COI) ≥ 1.0, and C) DiaSorin: positive cut-off ≥ 15 AU/mL and equivocal 12-15 AU/mL. In C negative samples with signals below detection limit (3.8 AU/ml) were plotted as 1.9 AU/mL.

The overall agreement was 97.5% for Abbott, 98.3% for Roche, 94.0% for DiaSorin, and 94.2% for Dynamiker Biotechnology. The pairwise inter-assay agreement (Cohen’s kappa) was as follows: Roche and Abbott 0.847 (95% CI, 0.718 to 0.976); Roche and Diasorin 0.654 (95% CI, 0.474 to 0.830); Roche and Dynamiker Biotechnology 0.794 (95% CI, 0.647 to 0.941); Abbott and DiaSorin 0.777 (95% CI, 0.618 to 0.936); Abbott and Dynamiker Biotechnology 0.804 (95% CI, 0.653 to 0.817); and DiaSorin and Dynamiker Biotechnology 0.724 (95% CI, 0.533 to 0.895).

The calculated sensitivity and specificity for each assay is presented in Table 2, together with the corresponding data extracted from each manufacturer’s test kit insert. In the present study Abbott exhibited the highest specificity and Roche the highest sensitivity, both being 100%. Two RT-PCR positive samples and two samples from the negative collection were excluded from the DiaSorin sample collection due to equivocal results. The positive likelihood ratios (LR+) were ∞, 50.0, 25.1 and 37.5 for Abbott, Roche, DiaSorin and Dynamiker Biotechnology, respectively.

**Table 2.**
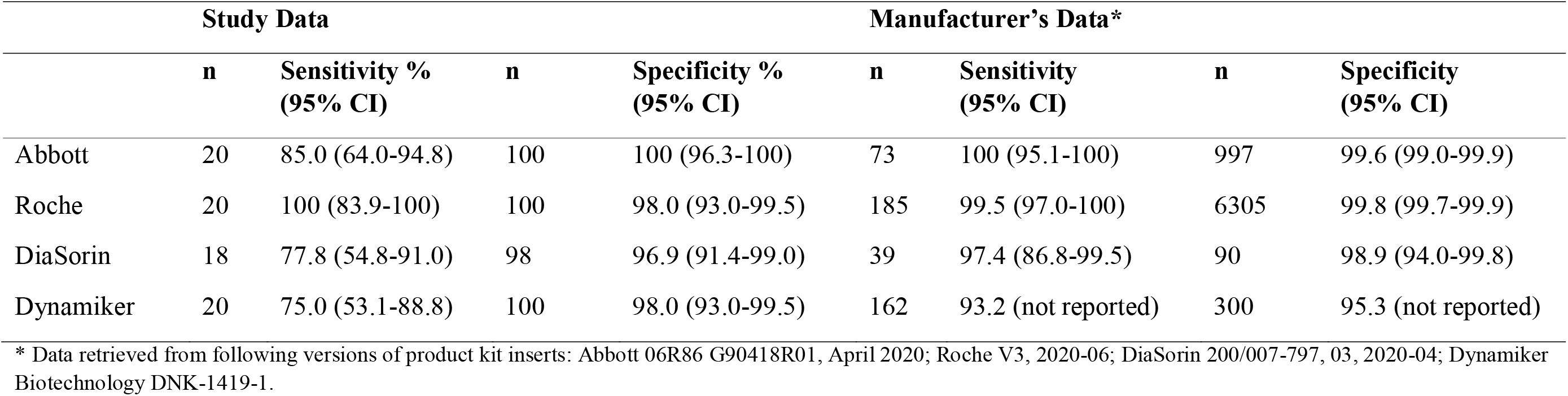
Sensitivity and specificity from this study, and according to the manufacturers’ data. The results are compared to manufacturers’ data from samples collected >14 days (Abbott, Roche) and >15 days (DiaSorin) post PCR confirmation. No information about time of sampling was available for Dynamiker Biotechnology.

Benchmarking the sensitivity and specificity data from the current study against published guidelines showed that the Abbott assay met the specificity criteria set by all three regulatory bodies. The specificities of Roche and Dynamiker Biotechnology were both in accordance with the HAS guidelines, while DiaSorin, in contrast, failed to reach any of the specificity criteria. Neither of the assays from Abbott, DiaSorin and Dynamiker Biotechnology exhibited a sufficient sensitivity to meet the criteria set by PHAS and HAS. In contrast, Roche performed excellent in this regard, with a sensitivity of 100%.

Assay performance recommendations from PHAS are based on a target PPV of > 90%. To demonstrate the seroprevalence required to reach a PPV of > 90%, the PPV based on sensitivity and specificity data from the present study as well as from the manufacturers, was calculated (Figure 2).

**Figure 2.**
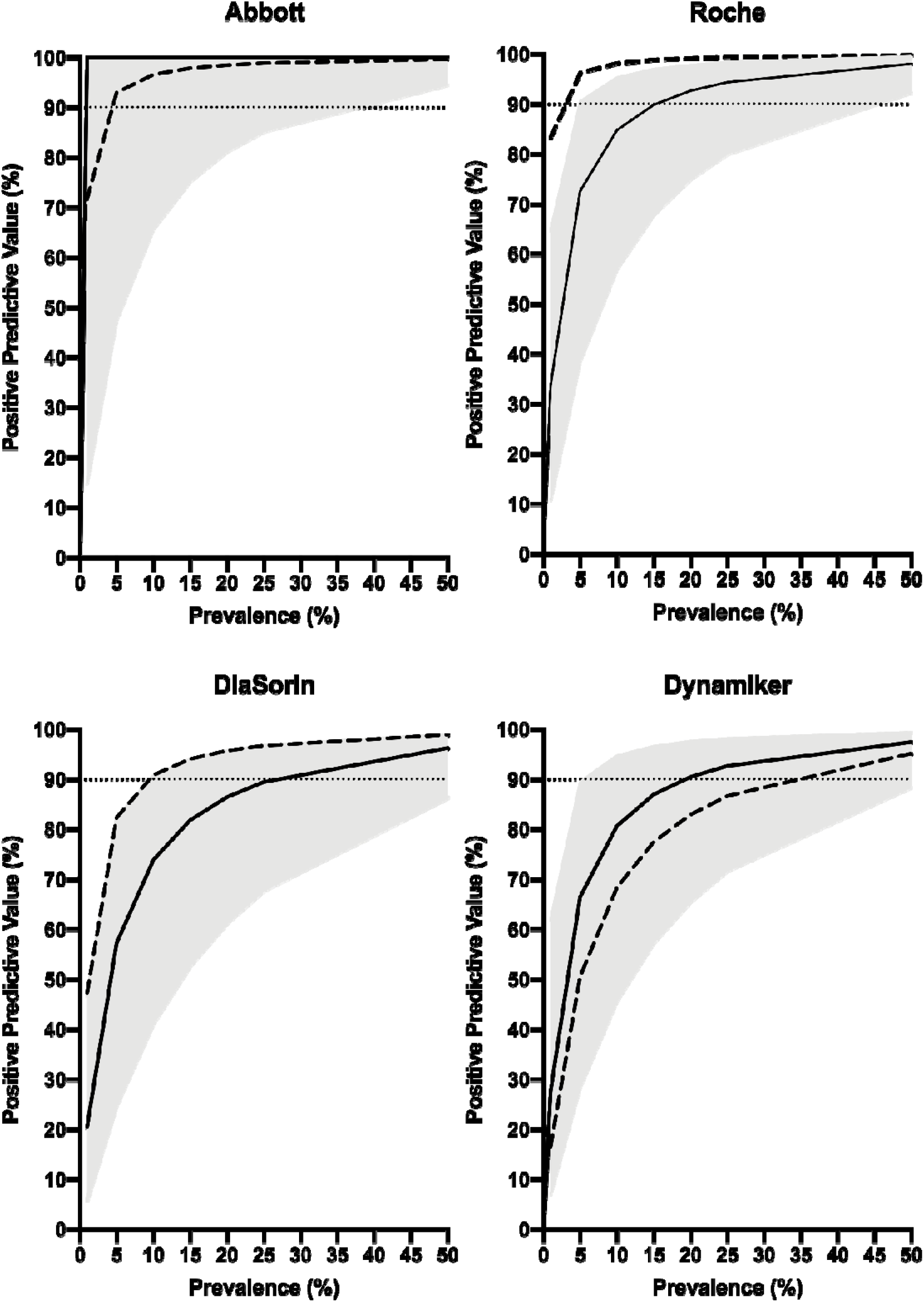
Estimated PPV for the depicted assays, calculated at seroprevalences of 1%, 5%, 10%, 15%, 20%, 25% and 50%. Calculations were based on sensitivity, specificity and their respective 95% CI limits from the current investigation (solid lines = mean values; grey areas = 95% CI). Dashed lines represent mean sensitivity and specificity data from the manufacturers’ kit inserts. Dotted horizontal lines refer to 90% PPV.

## Discussion

As the pandemic progresses, it will likely become increasingly important not only to establish ongoing COVID-19 but also to confirm past infection of SARS-CoV-2. On a community level, this could be used for assessing the progress of the pandemic and for guiding public health and control policies [10]. On an individual level, a reliable method for proving past COVID-19 could potentially be important for allowing employees to remain at work despite mild respiratory symptoms. It could even serve as a basis for a proposed “immunity passport” [11, 12]. Although the degree of immunity to SARS-CoV-2 is disputed, it is plausible that individuals undergoing COVID-19 will gain partial or temporary protection against new episodes [13]. Serology tests might also be used to identify donors of convalescent plasma, which has been proposed for the treatment of seriously ill COVID-19 patients [14].

However, the rapidly emerging pandemic limits the possibility of state-of-the-art validations according to the EP-12 A2 [6], and there is no international consensus on clinical performance requirements for COVID-19 assays. Neither is there an agreed gold standard method for antibody testing [15]. Using the performance criteria proposed by PHAS, HAS and CDC [7-9], only Abbott managed to reach the specificity recommended by all three authorities while Roche alone fulfilled the sensitivity criteria (PHAS, HAS).

The clinical performance of a qualitative diagnostic test is, with few exceptions, heavily dependent on the prevalence. Both PHAS and CDC accordingly link their respective specificity criteria for SARS-CoV-2 antibody testing to seroprevalence levels. PHAS recommends that a PPV > 90% shall be pursued, regardless of seroprevalence. Based on our results prevalences of approximately 15%, 20% and 25% would be necessary to reach this goal for Roche, Dynamiker Biotechnology and DiaSorin, respectively (Figure 2). In contrast, Abbott would in this regard perform well at any given seroprevalence, owing to its specificity of 100%.

Due to the limited sample size of the study, our results need to be interpreted with caution. Nevertheless, a recent study based on 65 RT-PCR positive and 1154 pre-COVID-19 samples [16] reported similar assay performances, except for the sensitivity of Roche being higher in our study. In contrast, evaluating four different assays using the same outpatient sample collection strengthens this study. Moreover, an aspect not investigated in this study is that assay performance could potentially vary with sample type (serum, plasma, whole blood).

Judging by the results from this study, introducing broad antibody testing for COVID-19 will be cumbersome in a low-prevalence setting. In order to reach the proposed performance criteria, two-tier testing will have to be considered. Alternatively, a high degree of specificity (and thus a high positive predictive value) could be prioritised using modified cut-off values. Our results suggest that this could be conceivable for the Abbott and Roche assays, while the more narrowly distributed results for DiaSorin would make it difficult to find such a specific cut-off with a reasonably preserved degree of sensitivity (Figure 1).

Considering the novelty of the disease, and hence the expedited development of new assays, possible sources of systematic errors (e.g. cross-reacting antibodies against other *Coronaviridae*) need to be carefully considered. In this study, all eleven false-positive samples originated from unique donors, none of the samples being positive in more than one assay. Further investigation of possible reasons for the false-positive results is, however, beyond the scope of this evaluation.

We conclude that the SARS-CoV-2 assays only partly fulfil the performance criteria proposed by regulatory authorities. Introduction into clinical use in low-prevalent settings, must therefore, be made with careful consideration and well-informed stakeholders.

## Data Availability

The data are available from the corresponding author on a reasonable request.

## Acknowledgements

The expert technical assistance of Ola Forsell and Susanna Bergqvist at the Department of Clinical Chemistry and Transfusion Medicine, Växjö Central Hospital, and of Sanna Hjalmarsson, Christina Bojesson and Eline Boesen at the Department of Clinical Microbiology, Region Kronoberg, is greatly appreciated.

## References

[1] N. Zhu, D. Zhang, W. Wang, X. Li, B. Yang, J. Song, X. Zhao, B. Huang, W. Shi, R. Lu, P. Niu, F. Zhan, X. Ma, D. Wang, W. Xu, G. Wu, G.F. Gao, W. Tan, I. China Novel Coronavirus, T. Research, A Novel Coronavirus from Patients with Pneumonia in China, 2019, N Engl J Med 382(8) (2020) 727–733.

[2] A.E. Gorbalenya, S.C. Baker, R.S. Baric, R.J. De Groot, C. Drosten, A.A. Gulyaeva, B.L. Haagmans, C. Lauber, A.M. Leontovich, B.W. Neuman, D. Penzar, S. Perlman, L.L.M. Poon, D. Samborskiy, I.A. Sidorov, I. Sola, J. Ziebuhr, Severe acute respiratory syndrome-related coronavirus: The species and its viruses - a statement of the Coronavirus Study Group, Cold Spring Harbor Laboratory, 2020.

[3] WHO, Virtual press conference on COVID-19-11 March 2020. <https://www.who.int/docs/default-source/coronaviruse/transcripts/who-audio-emergencies-coronavirus-press-conference-full-and-final-11mar2020.pdf?sfvrsn=cb432bb3_2>, 2020 (accessed 6th May.2020).

[4] Z. Zainol Rashid, S.N. Othman, M.N. Abdul Samat, U.K. Ali, K.K. Wong, Diagnostic performance of COVID-19 serology assays, Malays J Pathol 42(1) (2020) 13–21.

[5] V.M. Corman, O. Landt, M. Kaiser, R. Molenkamp, A. Meijer, D.K. Chu, T. Bleicker, S. Brunink, J. Schneider, M.L. Schmidt, D.G. Mulders, B.L. Haagmans, B. van der Veer, S. van den Brink, L. Wijsman, G. Goderski, J.L. Romette, J. Ellis, M. Zambon, M. Peiris, H. Goossens, C. Reusken, M.P. Koopmans, C. Drosten, Detection of 2019 novel coronavirus (2019-nCoV) by real-time RT-PCR, Euro Surveill 25(3) (2020).

[6] P.E. Garrett, F.D. Lasky, K.L. Meier, L.W. Clark, Clinical and Laboratory Standards Institute., User protocol for evaluation of qualitative test performance: approved guideline, 2nd ed., Clinical and Laboratory Standards Institute, Wayne, Pa., 2008.

[7] Folkhälsomyndigheten (Public Health Agency of Sweden), Vägledning för antikroppspåvisning. <https://www.folkhalsomyndigheten.se/contentassets/2c3d8e40926e4bcc942aa640922bb758/vagledning-antikroppspavisning.pdf>, 2020 (accessed 3 Jul.2020).

[8] Haute Authorité de Santé, Specifications setting out the performance assessment methods applicable to serological tests detecting anti-SARS-CoV-2 antibodies. <https://www.hassante.fr/upload/docs/application/pdf/2020-05/has_serological_tests_covid19_specifications.pdf>, 2020 (accessed Jul 31.2020).

[9] Centers for Disease Control and Prevention, Interim Guidelines for COVID-19 Antibody Testing; Interim Guidelines for COVID-19 Antibody Testing in Clinical and Public Health Settings. <https://www.cdc.gov/coronavirus/2019-ncov/lab/resources/antibody-tests-guidelines.html>, 2020 (accessed Jul 31.2020).

[10] S.E.F. Yong, D.E. Anderson, W.E. Wei, J. Pang, W.N. Chia, C.W. Tan, Y.L. Teoh, P. Rajendram, M. Toh, C. Poh, V.T.J. Koh, J. Lum, N.M. Suhaimi, P.Y. Chia, M.I. Chen, S. Vasoo, B. Ong, Y.S. Leo, L. Wang, V.J.M. Lee, Connecting clusters of COVID-19: an epidemiological and serological investigation, Lancet Infect Dis (2020).

[11] A.L. Phelan, COVID-19 immunity passports and vaccination certificates: scientific, equitable, and legal challenges, Lancet (2020).

[12] N.J. Beeching, T.E. Fletcher, M.B.J. Beadsworth, Covid-19: testing times, BMJ 369 (2020) m1403.

[13] L. Bao, W. Deng, H. Gao, C. Xiao, J. Liu, J. Xue, Q. Lv, J. Liu, P. Yu, Y. Xu, F. Qi, Y. Qu, F. Li, Z. Xiang, H. Yu, S. Gong, M. Liu, G. Wang, S. Wang, Z. Song, W. Zhao, Y. Han, L. Zhao, X. Liu, Q. Wei, C. Qin, Reinfection could not occur in SARS-CoV-2 infected rhesus macaques, bioRxiv (2020) 2020.03.13.990226.

[14] R. Rubin, Testing an Old Therapy Against a New Disease: Convalescent Plasma for COVID-19, JAMA (2020).

[15] European Commission, Guidelines on COVID-19 in vitro diagnostic tests and their performance (2020/C 122 I/01) <https://eur-lex.europa.eu/legal-content/EN/TXT/PDF/?uri=QJ:C:2020:122I:FULL&from=SV>, 2020 (accessed Aug. 4.2020).

[16] T. Perkmann, N. Perkmann-Nagele, M.-K. Breyer, R. Breyer-Kohansal, O.C. Burghuber, S. Hartl, D. Aletaha, D. Sieghart, P. Quehenberger, R. Marculescu, P. Mucher, R. Strassl, O.F. Wagner, C.J. Binder, H. Haslacher, Side by side comparison of three fully automated SARS-CoV-2 antibody assays with a focus on specificity, medRxiv (2020) 2020.06.04.20117911.

